# Developing deep learning-based strategies to predict the risk of hepatocellular carcinoma among patients with nonalcoholic fatty liver disease from electronic health records

**DOI:** 10.1101/2023.11.17.23298691

**Authors:** Zhao Li, Lan Lan, Yujia Zhou, Ruoxing Li, Kenneth D. Chavin, Hua Xu, Liang Li, David J. H. Shih, W. Jim Zheng

**Author notes:** Corresponding Author: W. Jim Zheng, PhD, McWilliams School of Biomedical Informatics, University of Texas Health Science Center at Houston, 7000 Fannin St, Houston, TX 77030, USA.

## Abstract

**Background:** Deep learning models showed great success and potential when applied to many biomedical problems. However, the accuracy of deep learning models for many disease prediction problems is affected by time-varying covariates, rare incidence, and covariate imbalance when using structured electronic health records data. The situation is further exasperated when predicting the risk of one disease on condition of another disease, such as the hepatocellular carcinoma risk among patients with nonalcoholic fatty liver disease due to slow, chronic progression, the scarce of data with both disease conditions and the sex bias of the diseases.

**Objective:** The goal of this study is to investigate the extent to which time-varying covariates, rare incidence, and covariate imbalance influence deep learning performance, and then devised strategies to tackle these challenges. These strategies were applied to improve hepatocellular carcinoma risk prediction among patients with nonalcoholic fatty liver disease.

**Methods:** We evaluated two representative deep learning models in the task of predicting the occurrence of hepatocellular carcinoma in a cohort of patients with nonalcoholic fatty liver disease (n = 220,838) from a national EHR database. The disease prediction task was carefully formulated as a classification problem while taking censorship and the length of follow-up into consideration.

**Results:** We developed a novel backward masking scheme to evaluate how the length of longitudinal information after the index date affects disease prediction. We observed that modeling time-varying covariates improved the performance of the algorithms and transfer learning mitigated reduced performance caused by the lack of data. In addition, covariate imbalance, such as sex bias in data impaired performance. Deep learning models trained on one sex and evaluated in the other sex showed reduced performance, indicating the importance of assessing covariate imbalance while preparing data for model training.

**Conclusions:** Devising proper strategies to address challenges from time-varying covariates, lack of data, and covariate imbalance can be key to counteracting data bias and accurately predicting disease occurrence using deep learning models. The novel strategies developed in this work can significantly improve the performance of hepatocellular carcinoma risk prediction among patients with nonalcoholic fatty liver disease. Furthermore, our novel strategies can be generalized to apply to other disease risk predictions using structured electronic health records, especially for disease risks on condition of another disease.

## Introduction

Hepatocellular carcinoma (HCC) is one of the most common types of primary liver cancer in adults and one of the leading causes of cancer-related deaths worldwide [1, 2]. Besides the well-known risk factors associated with HCC, e.g., hepatitis C, hepatitis B and alcoholic cirrhosis, nonalcoholic fatty liver disease (NAFLD) has also been linked to HCC in the United States and worldwide [3]. The previous analysis identified non-alcoholic steatohepatitis accounting for the underlying etiology in a small-scale analysis [4] and recognized sex as an important factor. However, these previous studies were relatively small with very limited number of patients who developed HCC after diagnosis of the liver diseases.

The extensive deployment of electronic health records (EHR) systems in the United States has accumulated vast amounts of patient medical history data [5]. Meanwhile, artificial intelligence, especially deep learning has shown great promise in clinical informatics research at large scale. Various deep learning approaches have been employed to predict HCC risk using clinical data. Ioannou et al. [6] applied Recurrent Neural Network to predict the HCC development among patients with hepatitis C virus–related cirrhosis in the national Veterans Health Administration database. Similarly, researchers also employed Convolutional Neural Network-based models to predict HCC development among viral hepatitis patients and patients with cirrhosis [7, 8]. These studies showed deep learning methods can achieve superior performance over the conventional regression models but none of them applied deep learning for predicting HCC development in patients with NAFLD [9–11]. Therefore, applying the latest deep learning approaches to analyze EHR data from large cohorts of patients for accurate assessment of the risk of developing HCC in patients with NAFLD is desirable.

Machine learning models [12, 13], and more recently, deep learning models [14–16], are used for disease prediction on structured data from EHR. Despite their great promise and improvement in performance, several inherent data challenges persist in applying deep learning-based approaches for disease risk prediction. For example, censoring occurs when certain data points within a study are either incomplete or unknown due to factors such as being outside the study’s time frame or scope, or other limitations. Censored data is very common in clinical, especially EHR-based research, because many patients will lose follow-up during the entire time frame of the clinical event of interest [17–19]. Current deep learning models often do not account for censoring in time-to-event data or have ad hoc design [19]. However, not properly accounting for censorship would lead to biased estimates of disease risk [20]. To utilize well-established machine learning algorithms for classification while handling censorship in time-to-event data, Craig et al. [21, 22] proposed to stack the features and outcomes of survival data at each timepoint into a single large table where the event time is cast as an additional covariate. However, this approach would lead to significant computing challenge for large datasets that are very common in EHR data. Additionally, the slow, chronic progression of diseases poses another challenge for disease risk prediction, as delayed diagnosis makes it difficult to predict future disease [23]. Time-varying covariates contain longitudinal information that could be important in disease risk prediction but have not been thoroughly evaluated while applying deep learning-based approaches [24–26]. Furthermore, deep learning-based approaches rely on large amounts of data for good performance, but predicting the risk of a disease in the context of another pre-existing disease typically results in small cohorts, as patients must satisfy the selection criteria for both diseases. To tackle this issue of data insufficiency, clinical concepts embeddings generated from a large EHR dataset were imported as initial embeddings based on other disease prediction tasks [27]. A transformer-based model was pretrained by masked language modeling on a large EHR database and can be finetuned to downstream tasks with small sample sizes [28]. However, the improvement of using these pretrained embeddings or models depends largely on the relatedness of the learning task and data during the pretraining stage and could be limited if the training data is dissimilar to the target data [29]. Besides these methodologic considerations, accurate predictions from machine learning models also strongly depend on the quality of input features and the training set [30]. Machine learning models may generate inferior or unreliable predictions if the unlabeled samples have dissimilar characteristics compared with the training set.

In this study, we investigate the impact of these challenges on performance of deep learning models in predicting disease risks from structured EHR data. We focus on hepatocellular carcinoma (HCC), one of the most common types of primary liver cancer in adults and one of the leading causes of cancer-related deaths worldwide [1, 2]. We predicted HCC risk among a large retrospective cohort of NAFLD patients (47% male and 53% female) from an EHR database containing records for over 68 million patients in the U.S. We formulated disease prediction as a classification problem while accounting for censoring and developed a novel approach to address delayed diagnosis by masking data before the diagnosis of disease. Our results demonstrate that time-varying covariate is a key factor influencing predictive performance. In addition, we established a new transfer learning paradigm for deep learning-based disease prediction on EHR data. Finally, we evaluated the impact of sex bias on deep learning performance and identified sex-specific features for HCC progression. Our study offers several key contributions. Firstly, we propose new approaches to preprocess event-to-event data from electronic health records (EHRs), which can improve the accuracy of downstream machine learning models. Secondly, we identify biases in EHR data that were previously overlooked or neglected, highlighting the importance of careful data preprocessing in medical research. Finally, we propose a novel strategy, backward masking, to deal with the issue of delayed diagnosis which is very common in EHR data analysis. Given the abundance of existing machine learning models and the vast amounts of EHR data available, we believe that addressing the challenges associated with applying machine learning to EHRs has broader implications for related fields as well.

## Methods

### Study Cohorts

The data in this study were extracted from the Cerner Health Facts® database, which contains deidentified EHR data for more than 68 million patients for the years 2000 to 2017. We extracted all encounter records for patients who satisfied the inclusion and exclusion criteria for each patient set described below. These encounter records contain data about demographics, diagnoses, medications, lab tests, and clinical events. Since Cerner Health Facts® used both International Classification of Diseases version 9 and version 10 for patient diagnoses, we standardized the medical codes by converting all version 9 codes to version 10 codes. We used the generic name for medications, and we used LOINC codes for lab tests and clinical events. Using the Cerner Health Facts® database, we created a set of patients with NAFLD and a case-control set. **Figure 1** shows the study flowchart.

**Figure 1.**
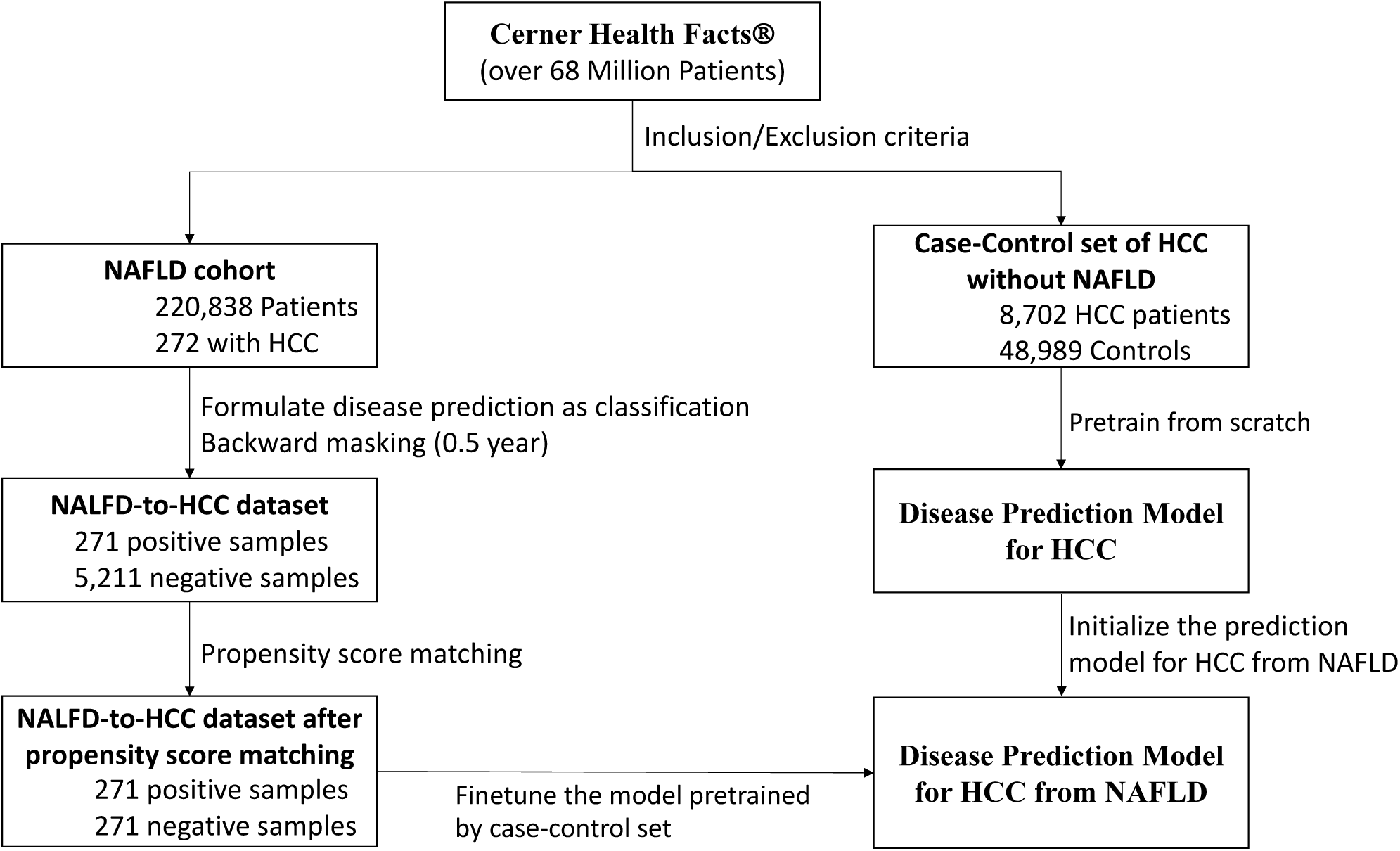
Study flowchart. We only show the backward masking of 0.5 year here for the purpose of illustration. Propensity score matching was conducted for all periods of backward masking.

### NAFLD cohort

Patients in this cohort had at least two abnormal alanine aminotransferase values over a > 6-month period [31]. Abnormal alanine aminotransferase values exceed 40 IU/mL for men and 31 IU/mL for women. We defined the date of the first abnormal alanine aminotransferase test as the index date. Patients who were younger than 18 years old on the index date were excluded. To preserve sufficient follow-up and quality of the included data, we followed the inclusion and exclusion criteria used by Kanwal *et al* [31], and we thus excluded those patients with less than 5 years of data in the Cerner Health Facts® database after the index date of NAFLD. Since the data is used to evaluate both baseline and deep learning methods, any bias introduced by this exclusion criteria will have minimal impact on our method evaluation and is outweighed by the benefit of resulting high quality data as demonstrated by many studies [31–34]. We excluded patients with hepatitis B or hepatitis C virus to eliminate the impact of these well-known risk factors for HCC. Finally, we excluded patients with a history of alcoholism or chronic hepatitis before the index date (**Figure 1**).

### Case-control patient set

To increase the size of the NAFLD cohort for training a deep learning model to perform HCC prediction, we extracted all patients with HCC from the Cerner Health Facts® dataset and added a set of healthy patients as matched controls who were at least 18 years old at the first encounter and had not been diagnosed with NAFLD. A matching control patient was randomly sampled for each HCC patient at a ratio of 10:1 based on gender, age at the first encounter, and duration from the first to the last encounter (**Figure 1**).

The characteristics of the NAFLD cohort and the case-control set are summarized in **Table 1**.

**Table 1.**
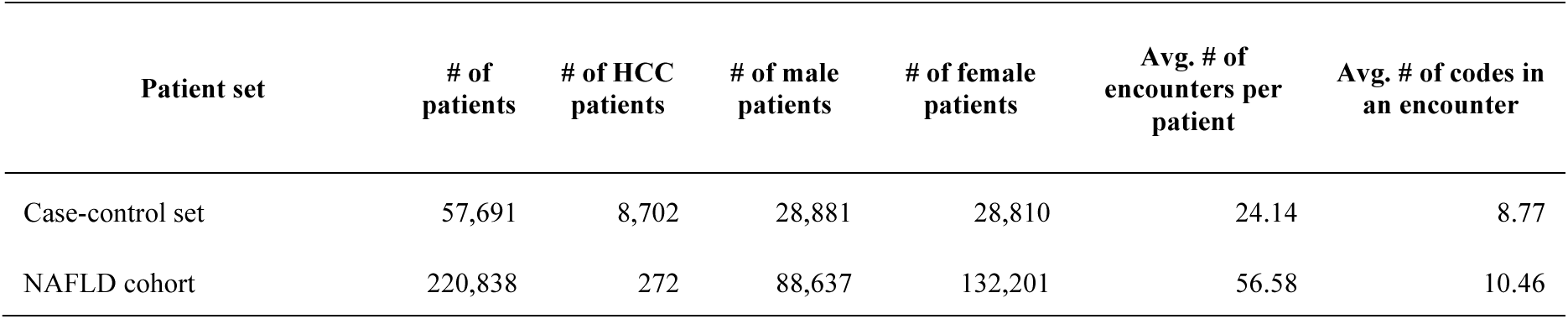
Summary characteristics of patient sets.

| Patient set | # of patients | # of HCC patients | # of male patients | # of female patients | Avg. # of encounters per patient | Avg. # of codes in an encounter |
| --- | --- | --- | --- | --- | --- | --- |
| Case-control set | 57,691 | 8,702 | 28,881 | 28,810 | 24.14 | 8.77 |
| NAFLD cohort | 220,838 | 272 | 88,637 | 132,201 | 56.58 | 10.46 |

### Study design

Deep learning models are usually designed for classification, and often do not account for censoring. We are interested in applying state-of-the-art deep learning models designed for classification to identify risk factors, rather than to estimate absolute risk precisely. However, not properly accounting for censorship can lead to biased estimates of disease risk [20]. Therefore, we carefully formulated disease prediction as a classification problem while considering censorship and the length of follow-up [22, 35].

We defined the event as the occurrence of HCC within 10 years after the index date using four individual patients as examples (**Figure 2**). Patients lost to follow-up for any reason, including death, were considered right-censored. We selected and labeled patients as shown in **Figure 2**. Patients who developed HCC after 10 years after index date were labeled as negative (Patient 1); those who did not develop HCC after the index date and were still in the Cerner Health Facts® database more than 10 years after the index date were also labeled as negative (Patient 2); the remaining patients were labeled as positive if they developed HCC within 10 years after index date (Patient 3); and all other patients were excluded (Patient 4). This last group of patients includes those who did not develop HCC but had less than 10 years of follow-up after the index date. Since there is insufficient information about whether these patients had developed HCC within 10 years of the index date, they do not have clear labels about their disease status at this time point, and they are thus not informative [17]. Under our criteria, patients who developed HCC before NAFLD were excluded, because we are interested in progression from NAFLD to HCC. Moreover, patients who developed HCC within 2 years after the index date were also excluded, since we focused on progression from NAFLD to HCC. If HCC is developed too quickly after NAFLD diagnosis, that means the patient might have HCC at the time NAFLD was diagnosed. This exclusion criterion might help the deep learning model pick up more meaningful predictive features for HCC prediction. Hence, the classification task for deep learning models is to estimate whether a patient with NAFLD will develop HCC within 10 years.

**Figure 2.**
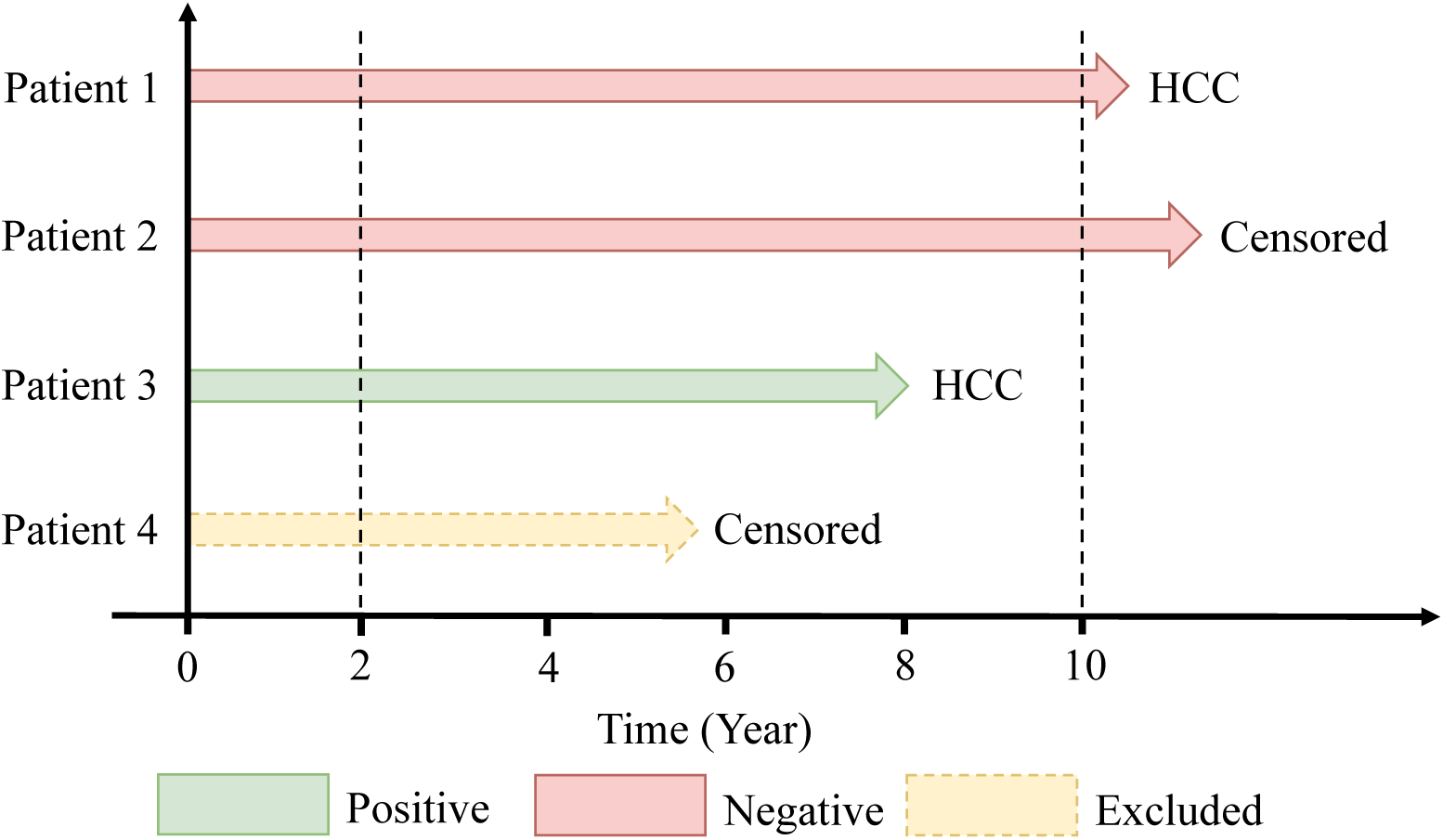
Study design. Formulation of disease prediction as a classification problem while accounting for censoring. Patients were included only if they had at least one year of medical history (dashed line at left) in the database. Samples were filtered and labeled as shown.

After the labeling process and backward masking (described below), we conducted propensity score matching to balance the ratio of positive and negative samples using the “MatchIt” R package. Features for conducting propensity score matching contain the mean and standard deviation of the time intervals among two successive encounters and the number of encounters for a patient. For each positive sample, we selected one control by performing greedy nearest-neighbor matching (**Figure 1**). The **Figure S1** shows the distribution of negative samples is closer to the positive samples after propensity score matching.

### Deep learning models and features

We used the two most representative deep learning models for our study: RETAIN is a classification-based model [24] and DeepHit is a deep survival model [25, 26]. RETAIN is a recurrent neural network-based model that uses an attention mechanism [24]. The gated recurrent unit inside RETAIN can efficiently use longitudinal medical information to predict disease progression. Meanwhile, a two-level attention module on top of the gated recurrent unit layers can generate the contribution weights of each feature (medical code) to the final prediction. Specifically, RETAIN first represents each medical code that is recorded in an encounter with a fixed-size, randomly initialized vector. All medical codes in an encounter are summed together to represent this encounter. Next, two gated recurrent unit layers with attention are used to generate the encounter-level and variable-level attention weights by the softmax function and hyperbolic tangent function, respectively. Finally, RETAIN represents each patient as a context vector that is a summation of all encounters, weighted by encounter-level and variable-level attention. Additionally, to mimic clinical practice, RETAIN takes the input of the EHR data in a reverse chronological order so that the model pays more attention to more recent encounters. A fully connected layer with the softmax activation function is used to calculate the final probability of the class labels, in this study HCC or no HCC. **Figure S2** shows the overview of RETAIN model. The input of RETAIN model contains the following features in all the included encounters: demographic (age, gender, race, and marital status), diagnosis codes (ICD 9 and ICD 10 code), medications in the format of the generic medication name, lab test and clinical event (LOINC code).

Besides RETAIN, we also used a deep learning survival model, DeepHit [25, 26], to predict survival times while taking into account the competing risk of death. DeepHit uses the baseline information of the index date and employed a deep neural network to learn the relationship between the input variables and the distribution of the survival times. Furthermore, DeepHit can handle the competing risk by incorporating cause-specific sub-networks for each event and a shared sub-network to learn the feature representations. The loss function in DeepHit includes the log-likelihood of the joint distribution of the first hitting time and corresponding event. Contrary to our application of RETAIN, we did not pre-process input data for censoring here because DeepHit handles censorship explicitly. However, while RETAIN uses the full longitudinal information of each patient for disease prediction, DeepHit only uses baseline information at the time of diagnosis of NAFLD. The complete list of baseline features is shown in **Table S1** in Supplementary Materials. Since DeepHit predicted the risk of HCC at each time point after the index date, to compare DeepHit to RETAIN, we transformed DeepHit outputs into the risk probability for HCC at 10 years after the index date. The Area Under the Receiver Operating Characteristic Curve (AUC) was the evaluation metric to benchmark these two methods [24, 36]. DeepHit and RETAIN are representative of two main categories of deep learning-based methods for disease prediction on EHR data: classification-based models that can use time-varying covariates to achieve more accurate predictions but do not properly account for censorship, which would lead to biased estimates of disease risk; deep survival models handle censorship properly but lack the capability to utilize time-varying covariates. In this analysis, we are not directly comparing DeepHit and RETAIN, but rather exploring whether modeling time-varying covariates with an unknown time-dependent function can lead to more accurate predictions.

### Backward masking

Due to the slow progression of HCC, some telltale signs and symptoms may be recorded in the medical record before a formal diagnosis code is recorded. However, we are interested in identifying risk factors, not well-known signs and symptoms of HCC. To mitigate the impact of delayed diagnosis, we trained multiple models by masking various lengths of medical history backward from the date of HCC diagnosis (**Figure 3**). This masking led to four subgroups, in which patient encounters within 0.5, 1, 2, or 4 years before HCC diagnosis or censoring are masked. We varied the masking length to evaluate how the duration of longitudinal information after the index date affects disease prediction. By masking different lengths of medical history of each patient in the NAFLD-to-HCC dataset, we obtained the training and testing data for each masking group to finetune and evaluate the model using five-fold cross validation.

**Figure 3.**
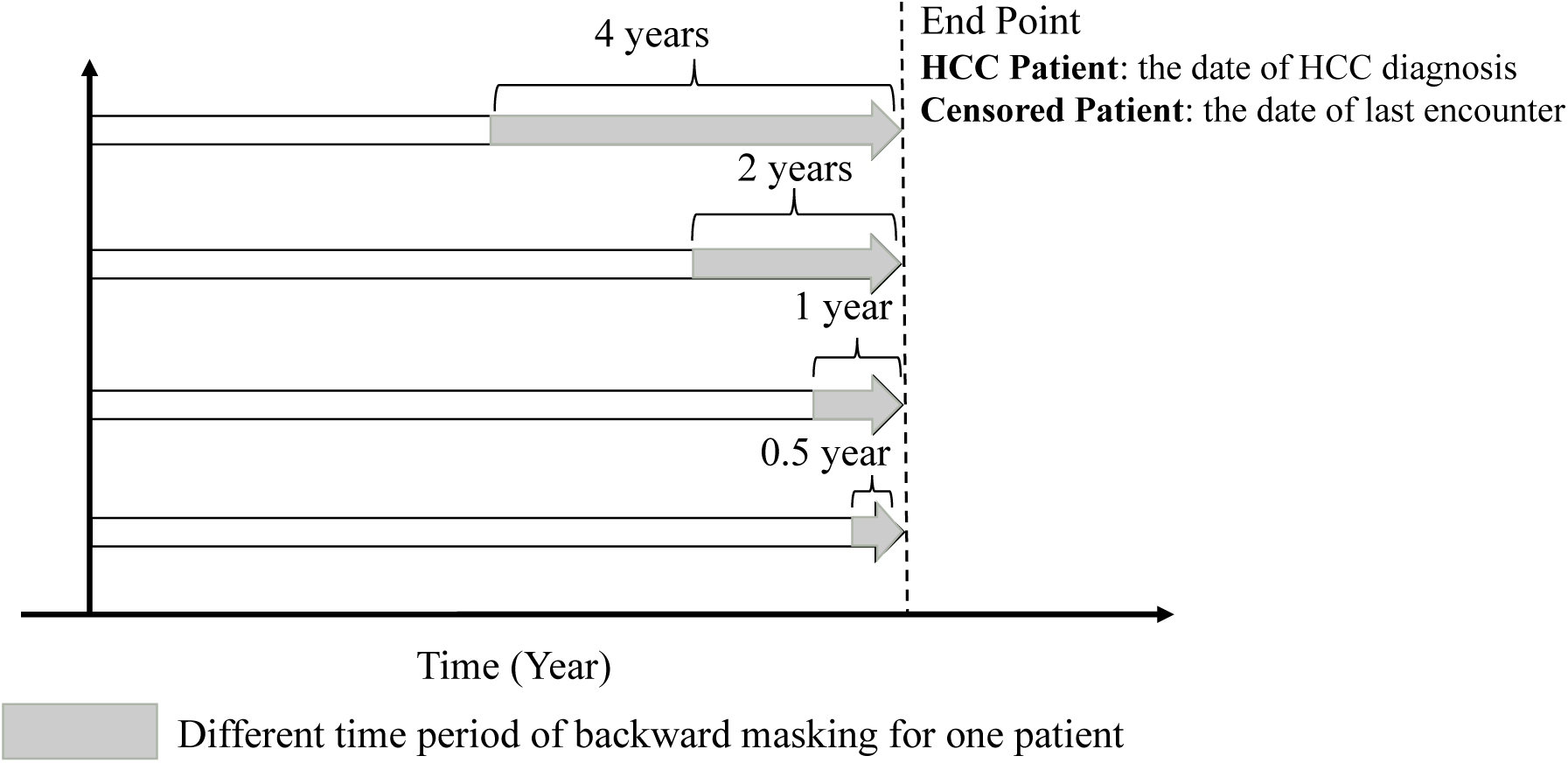
An example of masking various lengths of medical history backward from the date of HCC diagnosis or censoring. For HCC patients (Patient #1 and #3 in Figure 2), the end point is the date of HCC diagnosis. For censored patient without HCC (Patient #2 in Figure 2), the end point is the date of censoring, i.e. the last patient encounter in EHR. Patient #4 in Figure 2 was excluded.

### Transfer learning

As shown in **Table 1**, there are only 272 HCC patients in the NAFLD cohort. Since a typical deep learning model like RETAIN contains many learnable parameters, this sample would be inadequate for finding an acceptable solution in such a large parameter space. A common approach for circumventing small sample sizes in EHR data is to use a general-purpose pretrained model, and fine-tune the embedding layers of the network on the target dataset [13, 36]. In this study, we pretrained the whole model on a related prediction task for which a large dataset is available for training [27, 37, 38]. Specifically, we trained the deep learning on the larger case-control set, so that the model learns to recognize general patterns such as early symptoms and complications of HCC development. We then used this pretrained model and fine-tuned it on the NAFLD cohort, and we used cross-validation to evaluate the model’s performance in predicting HCC among NAFLD patients. We used the same dictionary of medical codes and model architecture during pretraining and finetuning.

### Aggregated attention scores

The attention scores outputted by the RETAIN model were employed to identify which factors contribute more to the prediction of HCC. Specifically, since RETAIN model output the attention scores for each code in all encounters and patients, we first calculated the mean of the attention score of each medical code across encounters for each patient to obtain the code-level importance score for this specific patient. Then we averaged the mean values of each code across different patients to obtain the importance value of each medical code within the cohort. Finally, we ranked the risk factors and protective factors by their cohort-level importance value and excluded medical codes observed in fewer than 10 patients. We applied this procedure to both male and female patients and identified sex-specific features for HCC progression.

## Results

### Formulating disease prediction as a classification problem

We compared the performance of two state-of-the-art deep learning algorithms for disease prediction: DeepHit and RETAIN. DeepHit incorporates a statistically rigorous competing risk model for fitting time-to-event data, while RETAIN is designed only for classification without consideration for censoring. However, DeepHit only considers covariate values at baseline, whereas RETAIN tracks changes in covariates across time. Since we are particularly interested in identifying risk factors and protective factors, rather than estimating absolute disease risk, we formulated disease prediction as a classification problem at a specified time point while accounting for censoring (Methods, **Figure 2**). We showed with Monte Carlo simulations that analyzing time-to-event data as a classification problem in this way allows us to identify risk factors reliably with strong control of type I error (Supplementary **Figure S3-5**).

### Modeling time-varying covariate improves disease prediction

We evaluated the predictive performance of RETAIN and DeepHit by five-fold cross-validation. Overall, RETAIN achieved a consistently high AUC score of ∼0.95 on all patients, male patients, and female patients, outperforming DeepHit (**Figure 4**). The error bars show the standard deviations of the AUC scores across 5 folds. When considering all patients or men only, RETAIN outperformed DeepHit by about 0.1 in AUC score. Among female patients, RETAIN outperformed DeepHit by ∼0.21 in AUC score. We hypothesized that RETAIN outperformed DeepHit because the RETAIN model used more longitudinal information. Henceforth, we focused on evaluating the performance characteristics of the much superior deep learning model, RETAIN.

**Figure 4.**
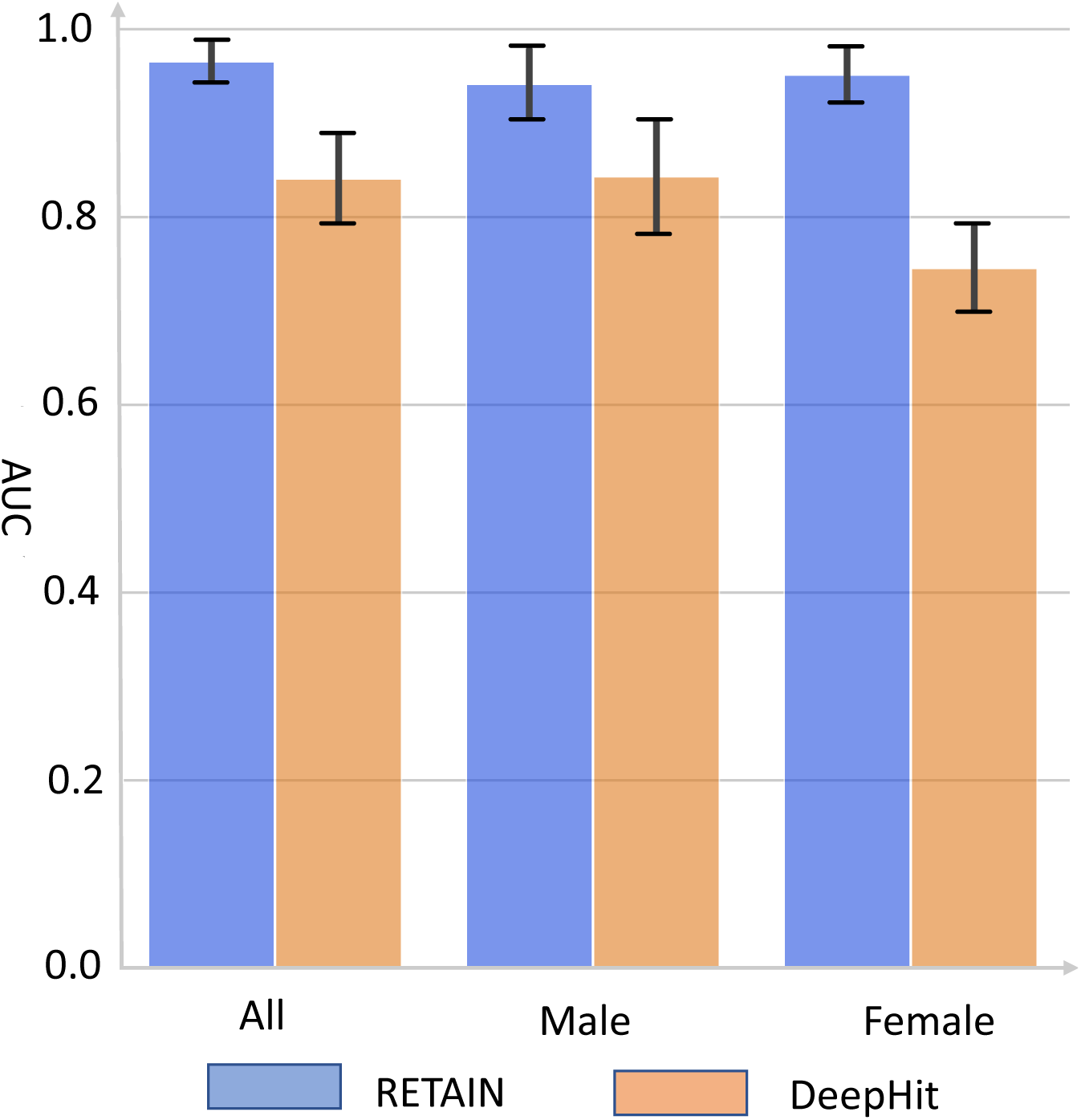
Predict HCC occurrence in NAFLD patients using RETAIN and DeepHit. All patients, male patients alone, or female patients alone were used respectively from the NAFLD cohort. Bars is the mean and error bars is the standard deviation of AUC from 5-fold cross- validation. The RETAIN model used covariate values from encounters up to 0.5 year before HCC diagnosis or censoring; DeepHit used only covariate values at baseline.

To test our hypothesis that time-varying covariates are critical to the superior predictive performance of RETAIN, we performed backward masking on the covariate data. That is, we masked data derived from encounters that occurred within 0.5, 1.0, 2.0, or 4.0 years before the date of disease diagnosis (or the final encounter for patients who never developed HCC). Hence, as we increased the masking length, more longitudinal information is withheld from the RETAIN model. With longer backward masking, the average length of an available patient’s medical history decreased. Concomitant with less longitudinal information (i.e. shorter medical history and fewer available encounters), the AUC score of RETAIN decreased from 0.966 to 0.906 for models evaluated with cross-validation on all patients (**Table 2**). Even larger performance reductions were observed for models trained and evaluated on only male or only female patients. The AUC decreased to 0.838 and 0.828 for men and women, respectively. With 4 years of backward masking, < 3 years of longitudinal data were available to RETAIN, which caused the AUC performance of RETAIN to decline to comparable levels as those achieved by DeepHit using only baseline covariates (**Table 2**, **Figure 4**). Taken together, these results indicate that RETAIN relies on the use of time-varying covariate values beyond baseline to achieve superior performance.

**Table 2.** The performance characteristics of RETAIN for different lengths of backward masking.

| *Masking length (yrs)* |  | *0.5* | *1.0* | *2.0* | *4.0* |
| --- | --- | --- | --- | --- | --- |
| *Medical history (yrs)* | Mean | 7.16 | 6.37 | 5.14 | 2.93 |
|  | Median | 7.27 | 6.43 | 5.05 | 2.84 |
| *Mean AUC (SD)* | All | 0.966(0.019) | 0.955(0.008) | 0.945(0.023) | 0.906(0.026) |
|  | Male | 0.942(0.036) | 0.908(0.035) | 0.914(0.046) | 0.838(0.057) |
|  | Female | 0.952(0.027) | 0.920(0.036) | 0.927(0.023) | 0.828(0.101) |

### Transfer learning improves model performance

We evaluated the extent to which transfer learning improved RETAIN performance. In our NAFLD cohort, only 2 of 1000 NAFLD patients developed HCC after 10 years beyond the index date. Due to the rare incidence of HCC among NAFLD patients, we pretrained RETAIN models on the larger set of patients who developed HCC (excluding NAFLD patients) and control patients who never developed HCC. We then fine-tuned these pretrained models on the NAFLD cohort. With transfer learning, AUC performances substantially increased, with improvements in AUC ranging from 0.019 to 0.095 across different lengths of backward masking (**Table 3**). The paired T-test was conducted to measure the statistical significance of using TL under different conditions in the AUC scores from 5 folds. The AUC after TL was subtracted from the AUC before TL, so the alternative hypothesis assumes that the mean difference of AUC after using TL is less than zero, which means the AUC after TL is larger than the AUC without TL. The improvement was statistically significant with Transfer Learning under many conditions. Transfer learning also reduced the validation loss (Supplementary **Figure S6**). These results indicate that RETAIN learned generalizable patterns from the larger case-control HCC dataset and that this information helped RETAIN achieved better performance on the smaller cohort of interest. Furthermore, greater improvements were observed with longer backward masking (**Table 3**). For example, the AUC improved 0.065 and 0.08 with less than 2 years masking for male and female patients, respectively. However, the improvements were just 0.019 and 0.038 with 0.5 year of backward masking. Moreover, models applied only in women achieved consistently higher improvement than models applied only in men, irrespective of masking length (**Table 3**).

**Table 3.**
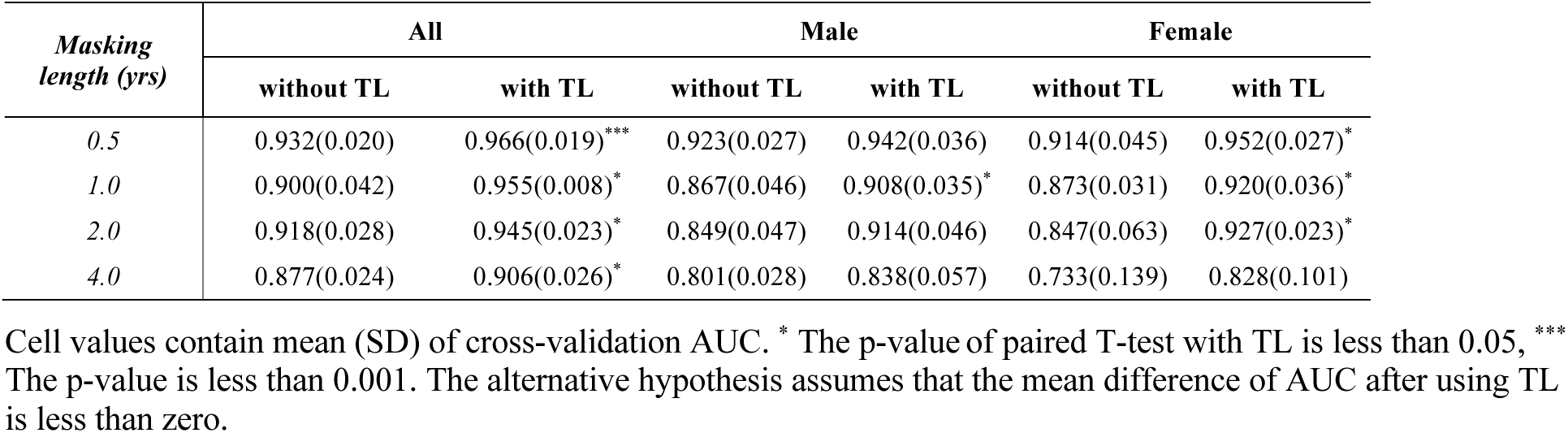
AUC performances of RETAIN models with and without transfer learning (TL).

### Sex bias impacts model performance

Gender disparity in HCC morbidity and survival outcome has been extensively studied and documented [10, 39–43]. However, Prior studies on HCC risk were either small [3, 4] or sex-biased where 94.4% of patients in the study were male [31], which may have led to biased results. Therefore, investigating sex bias in HCC can improve our understanding of how sex-based differences affect outcomes and help ensure data fairness in AI applications [40, 44]. To assess how sex bias in data affects the performance of deep learning, we trained RETAIN models on one sex and evaluated them in the other sex. As shown in **Table 4**, the model trained using male patients achieved inferior performance in female patients, as the AUC decreased from 0.927 to 0.834 with 2 years of backward masking. When applying the model trained on female patients with 4 years of backward masking, the performance for male prediction decreased from 0.838 to 0.773. These results are consistent with the concept that sex influences HCC risk, and there could be male- and female-specific features learned by the model that can only perform well in predicting HCC risks in that sex. As expected, the model trained on the data with both sexes achieved better performance than that trained on the data for either sex alone.

**Table 4.** The performance of RETAIN trained and evaluated on different subgroups. The scores are the average AUC (SD) from cross-validation.

| *Predicted group* | *Trained with* | *Masking length (years)* |  |  |  |
| --- | --- | --- | --- | --- | --- |
|  |  | 0.5 | 1 | 2 | 4 |
| *Male* | All* | 0.965(0.030) | 0.948(0.019) | 0.931(0.041) | 0.900(0.050) |
|  | Male | 0.942(0.036) | 0.908(0.035) | 0.914(0.046) | **0.838(0.057)** |
|  | Female | 0.890(0.059) | 0.848(0.051) | 0.873(0.046) | **0.773(0.065)** |
| *Female* | All | 0.964(0.018) | 0.964(0.014) | 0.960(0.026) | 0.916(0.009) |
|  | Male | 0.905(0.038) | 0.878(0.040) | **0.834(0.028)** | 0.806(0.086) |
|  | Female | 0.952(0.027) | 0.920(0.036) | **0.927(0.023)** | 0.828(0.101) |

\* Trained on both male and female patients.

### Sex-specific features

To understand why RETAIN achieves different predictive performances in male vs. female patients, we analyzed aggregated attention scores obtained from the model. As shown in **Table S2-S5** in Supplementary Materials, we identified medical codes with positive attention scores and negative attention scores for male and female patients. Some common and well-known risk factors or complications of HCC are ranked high in all patients, e.g., high body mass index, abnormal aspartate transferase values, and the presence of type 2 diabetes mellitus. Specifically for women, rheumatoid arthritis is associated with positive attention scores, and kidney stones are associated with negative attention scores.

To illustrate how RETAIN makes a specific prediction, in **Figure 5** we depict the attention scores of codes for a specific patient who developed HCC, which was correctly predicted. We filtered out the codes whose attention scores are between the 25^th^ percentile and the 75^th^ percentile in all encounters to make the figure readable. In the RETAIN prediction for this patient, normal lab results for tests such as platelet count and aspartate transferase were possible protective factors, whereas type 2 diabetes, hypertension, and elevated aspartate transferase were possible risk factors. Being a non-smoker was associated with a positive attention score.

**Figure 5.**
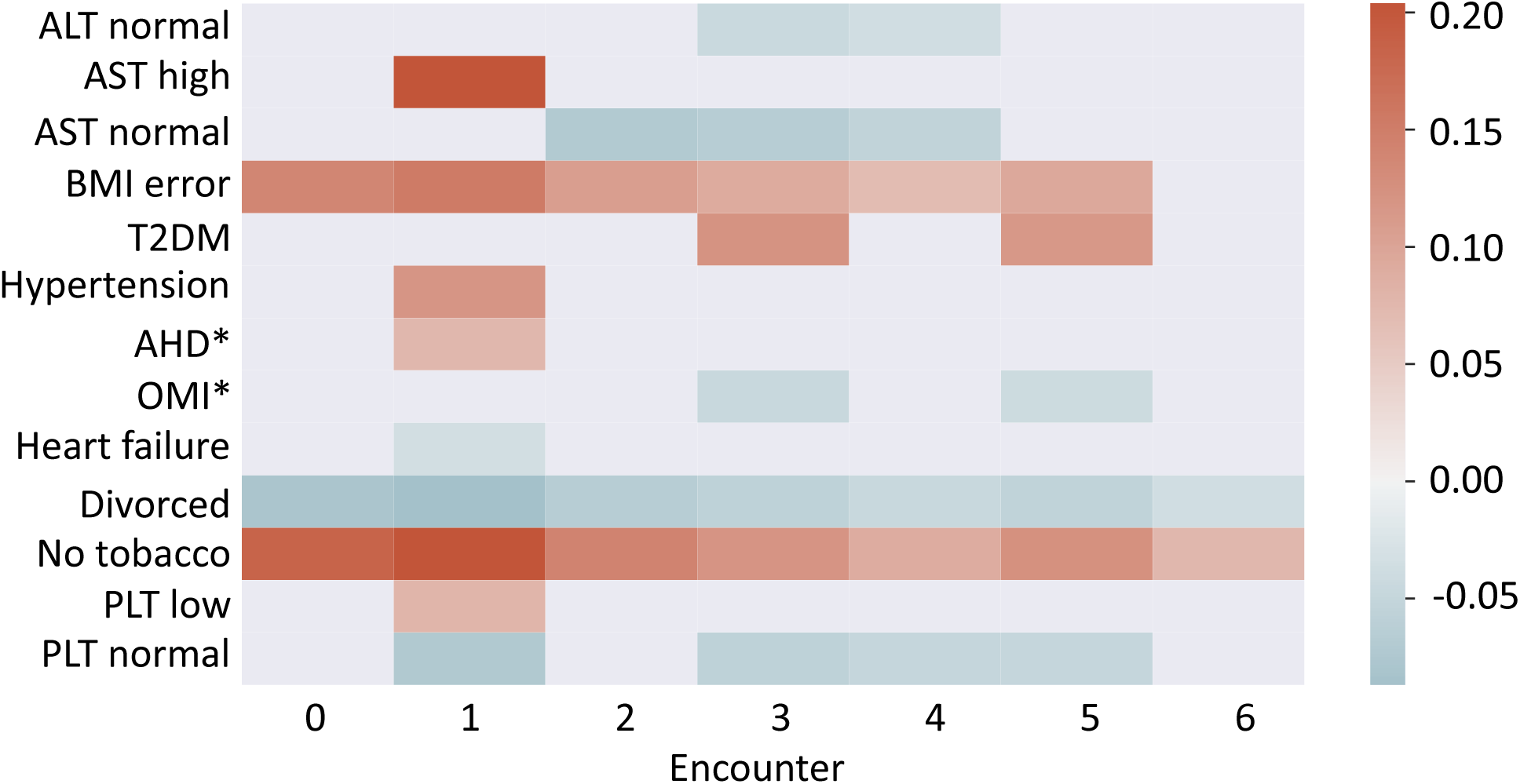
Visualization of RETAIN attention scores for a patient. Codes with attention scores higher than 75^th^ percentile or lower than 25^th^ percentile in at least one encounter are plotted. The color range represents the associated attention scores from the model. “BMI error” means the BMI value in the EHR database is invalid (negative or empty). AHD: Atherosclerotic Heart Disease. OMI: Old Myocardial Infarction.

## Discussion

We highlight key areas of innovation in our work for the application of deep learning in medical informatics. To compare deep learning algorithms using the same evaluation measure, we formulated disease prediction as a classification problem while taking censoring into account. To account for delayed disease diagnosis, we propose a backward masking approach, which prevents deep learning models that incorporate time-varying covariates from identifying telltale signs and symptoms of the disease rather than risk factors. Additionally, we showed that incorporating time-varying covariates with a deep learning model, such as RETAIN, can improve the AUC performance of the classification-based disease prediction. We then demonstrate that transfer learning improves predictive performance when the data set is limited by having insufficient number of cases. Further, we demonstrated that sex bias adversely impacts deep learning performance, and we identified sex-specific features for HCC progression.

Delayed diagnosis is common in clinical practice, and failure to take this issue into account may cause deep learning algorithms to identify trivial features that are not of interest to the clinical community. Patients have prescribed medications or diagnosed with symptoms for some diseases, but the actual entry of the diagnosis code for this disease can be delayed for quite some time due to factors including laboratory test confirmation and clerical errors. This delayed entry of the diagnosis code thus allows deep learning algorithms to predict disease status effortlessly using telltale symptoms or prescribed medications for the disease. To mitigate this issue, we designed a new masking strategy to investigate how delayed diagnosis impacts deep learning models that use longitudinal information. When we mask for a sufficient length of time before the formal diagnosis of the disease, we can prevent deep learning algorithms from using trivial features of the disease and allow the algorithms to identify more subtle patterns of features that help us gain critical clinical insights into the etiology and progression of the disease. Although some survival analyses can use the time-varying variates to make predictions at different time points (horizons) prior to death by manual feature selection, this is different from our design which aims to solve the issue of delayed diagnosis and telltale symptoms [45].

Using backward masking, we demonstrated that modeling time-varying covariate of features contributes strongly to disease prediction. When we masked patient encounters that occurred within four years of HCC diagnosis, the predictive performance of RETAIN (which models time-varying features) decreased to similar levels as that of DeepHit (which only models baseline features). This finding indicates that the improved disease prediction performance of RETAIN over DeepHit is due to the modeling of time-varying covariate of the features. Accordingly, incorporating time-varying covariates can be key to achieving optimal performance for disease prediction.

Furthermore, we showed that transfer learning can be used to remedy the rarity of positive examples. We recognized that despite beginning with a large cohort of millions of patients, the subset of patients who experienced the outcome event is relatively small. Different from the prior studies which typically adopt the embedding vectors of medical codes learned from a general task [27] or finetune a large pretrained model which was trained on diverse but unrelated medical conditions [28], we first trained a randomly initialized model on a related problem with large sample size, and finetuned the trained model on the target problem. We demonstrated that the application of this transfer learning strategy yielded a remarkable improvement in predictive performance. This improvement also indicates that although the larger, related dataset contains cases that are not of primary interest, they may share general patterns that help deep learning algorithms discern the disease of interest. Pretraining on the larger, related dataset thus improved prediction on the smaller target dataset.

## Data Availability

Since the Cerner database is commercial, anyone interested can subscribe to Cerner Health Facts to access the data.

## Acknowledgements

This work is partly supported by the National Institutes of Health (NIH) through grant 1UL1TR003167, 1R01AG066749-01, DoD W81XWH2210164 and the Cancer Prevention and Research Institute of Texas through grant RP170668 (WJZ). W.J.Z. conceived of the project. W.J.Z., D.J.H.S. and Z.L. designed and coordinated the study. Z.L., L. Lan, and Y.Z. preprocessed the data. Z.L. performed the deep learning model training and evaluation. R.L. performed the statistical simulations. K.D.C provided clinical advice. L. Li provided statistical guidance for the study. D.J.H.S., Z.L., L. Lan, and R.L. wrote the manuscript. All authors read and approved the final manuscript. We acknowledge the use of “Cerner Health Facts®” and the assistance provided by UTHealth SBMI Data Service team to extract data.

## Abbreviations

EHR: electronic health records
NAFLD: nonalcoholic fatty liver disease
HCC: hepatocellular carcinoma
AUC: Area Under the Receiver Operating Characteristic Curve

## References

1. 1. Forner A, Llovet JM, Bruix J: Hepatocellular carcinoma. The Lancet (British edition) 2012, 379(9822):1245–1255.

2. Venook AP, Papandreou C, Furuse J, Ladrón de Guevara L: The Incidence and Epidemiology of Hepatocellular Carcinoma: A Global and Regional Perspective. The oncologist (Dayton, Ohio) 2010, 15(S4):5–13.

3. Williams CD, Stengel J, Asike MI, Torres DM, Shaw J, Contreras M, Landt CL, Harrison SA: Prevalence of nonalcoholic fatty liver disease and nonalcoholic steatohepatitis among a largely middle-aged population utilizing ultrasound and liver biopsy: a prospective study. Gastroenterology 2011, 140(1):124–131.

4. Ertle J, Dechêne A, Sowa JP, Penndorf V, Herzer K, Kaiser G, Schlaak JF, Gerken G, Syn WK, Canbay A: Non-alcoholic fatty liver disease progresses to hepatocellular carcinoma in the absence of apparent cirrhosis. International journal of cancer 2011, 128(10):2436–2443.

5. Choi E, Bahadori MT, Schuetz A, Stewart WF, Sun J: Doctor AI: Predicting Clinical Events via Recurrent Neural Networks. JMLR workshop and conference proceedings 2016, 56:301–318.

6. Ioannou GN, Tang W, Beste LA, Tincopa MA, Su GL, Van T, Tapper EB, Singal AG, Zhu J, Waljee AK: Assessment of a deep learning model to predict hepatocellular carcinoma in patients with hepatitis C cirrhosis. JAMA network open 2020, 3(9):e2015626–e2015626.

7. Phan DV, Chan CL, Li AHA, Chien TY, Nguyen VC: Liver cancer prediction in a viral hepatitis cohort: A deep learning approach. International Journal of Cancer 2020, 147(10):2871–2878.

8. Nam JY, Sinn DH, Bae J, Jang ES, Kim J-W, Jeong S-H: Deep learning model for prediction of hepatocellular carcinoma in patients with HBV-related cirrhosis on antiviral therapy. JHEP Reports 2020, 2(6):100175.

9. Ahn JC, Qureshi TA, Singal AG, Li D, Yang J-D: Deep learning in hepatocellular carcinoma: Current status and future perspectives. World Journal of Hepatology 2021, 13(12):2039.

10. Huang FY, Wong DK, Seto WK, Lai CL, Yuen MF: Estradiol induces apoptosis via activation of miRNA-23a and p53: implication for gender difference in liver cancer development. Oncotarget 2015, 6(33):34941–34952.

11. Shah PA, Patil R, Harrison SA: NAFLD-related hepatocellular carcinoma: The growing challenge. Hepatology 2022.

12. Shickel B, Tighe PJ, Bihorac A, Rashidi P: Deep EHR: a survey of recent advances in deep learning techniques for electronic health record (EHR) analysis. IEEE journal of biomedical and health informatics 2017, 22(5):1589–1604.

13. Si Y, Du J, Li Z, Jiang X, Miller T, Wang F, Zheng WJ, Roberts K: Deep representation learning of patient data from Electronic Health Records (EHR): a systematic review. arXiv preprint arXiv:201002809 2020.

14. Tran T, Nguyen TD, Phung D, Venkatesh S: Learning vector representation of medical objects via EMR-driven nonnegative restricted Boltzmann machines (eNRBM). Journal of biomedical informatics 2015, 54:96–105.

15. Choi E, Bahadori MT, Schuetz A, Stewart WF, Sun J: Doctor ai: Predicting clinical events via recurrent neural networks. In: Machine Learning for Healthcare Conference: 2016. 301–318.

16. Choi E, Xu Z, Li Y, Dusenberry MW, Flores G, Xue Y, Dai AM: Graph Convolutional Transformer: Learning the Graphical Structure of Electronic Health Records. arXiv preprint arXiv:190604716 2019.

17. Klein JP, Moeschberger ML, collection EBe: Survival Analysis : Techniques for Censored and Truncated Data, 2nd 2003. edn. New York, NY: Springer New York : Imprint: Springer; 2003.

18. Schluchter MD, Greene T, Beck GJ: Analysis of change in the presence of informative censoring: application to a longitudinal clinical trial of progressive renal disease. Statistics in medicine 2001, 20(7):989–1007.

19. Vock DM, Wolfson J, Bandyopadhyay S, Adomavicius G, Johnson PE, Vazquez-Benitez G, O’Connor PJ: Adapting machine learning techniques to censored time-to-event health record data: A general-purpose approach using inverse probability of censoring weighting. Journal of biomedical informatics 2016, 61:119–131.

20. Katzman JL, Shaham U, Cloninger A, Bates J, Jiang T, Kluger Y: DeepSurv: personalized treatment recommender system using a Cox proportional hazards deep neural network. BMC medical research methodology 2018, 18(1):1–12.

21. Craig E, Zhong C, Tibshirani R: Survival stacking: casting survival analysis as a classification problem. arXiv preprint arXiv:210713480 2021.

22. Zhong C, Tibshirani R: Survival analysis as a classification problem. arXiv preprint arXiv:190911171 2019.

23. Ofosu A, Ramai D, Reddy M: Non-alcoholic fatty liver disease: controlling an emerging epidemic, challenges, and future directions. Ann Gastroenterol 2018, 31(3):288–295.

24. Choi E, Bahadori MT, Sun J, Kulas J, Schuetz A, Stewart W: Retain: An interpretable predictive model for healthcare using reverse time attention mechanism. In: Advances in Neural Information Processing Systems: 2016. 3504–3512.

25. Lee C, Yoon J, Van Der Schaar M: Dynamic-deephit: A deep learning approach for dynamic survival analysis with competing risks based on longitudinal data. IEEE Transactions on Biomedical Engineering 2019, 67(1):122–133.

26. Lee C, Zame W, Yoon J, van der Schaar M: Deephit: A deep learning approach to survival analysis with competing risks. In: Proceedings of the AAAI Conference on Artificial Intelligence: 2018.

27. Xiang Y, Xu J, Si Y, Li Z, Rasmy L, Zhou Y, Tiryaki F, Li F, Zhang Y, Wu Y: Time-sensitive clinical concept embeddings learned from large electronic health records. BMC medical informatics and decision making 2019, 19(2):139–148.

28. Rasmy L, Xiang Y, Xie Z, Tao C, Zhi D: Med-BERT: pretrained contextualized embeddings on large-scale structured electronic health records for disease prediction. NPJ digital medicine 2021, 4(1):1–13.

29. Xue Y, Du N, Mottram A, Seneviratne M, Dai AM: Learning to select best forecast tasks for clinical outcome prediction. Advances in Neural Information Processing Systems 2020, 33:15031–15041.

30. Murphy KP: Machine learning: a probabilistic perspective: MIT press; 2012.

31. Kanwal F, Kramer JR, Mapakshi S, Natarajan Y, Chayanupatkul M, Richardson PA, Li L, Desiderio R, Thrift AP, Asch SM et al: Risk of Hepatocellular Cancer in Patients With Non-Alcoholic Fatty Liver Disease. Gastroenterology 2018, 155(6):1828–1837.e1822.

32. Pittet D, Wyssa B, Herter-Clavel C, Kursteiner K, Vaucher J, Lew PD: Outcome of diabetic foot infections treated conservatively: a retrospective cohort study with long-term follow-up. Arch Intern Med 1999, 159(8):851–856.

33. White IK, Shaikh KA, Moore RJ, Bullis CL, Sami MT, Gianaris TJ, Fulkerson DH: Risk of radiation-induced malignancies from CT scanning in children who underwent shunt treatment before 6 years of age: a retrospective cohort study with a minimum 10-year follow-up. J Neurosurg Pediatr 2014, 13(5):514–519.

34. Yu Z, Chen J, Cheng X, Xie D, Chen Y, Zou X, Peng X: Patients with degenerative cervical myelopathy exhibit neurophysiological improvement upon extension and flexion: a retrospective cohort study with a minimum 1-year follow-up. BMC Neurol 2022, 22(1):110.

35. Ranganath R, Perotte A, Elhadad N, Blei D: Deep survival analysis. In: Machine Learning for Healthcare Conference: 2016. PMLR: 101–114.

36. Rasmy L, Wu Y, Wang N, Geng X, Zheng WJ, Wang F, Wu H, Xu H, Zhi D: A study of generalizability of recurrent neural network-based predictive models for heart failure onset risk using a large and heterogeneous EHR data set. Journal of biomedical informatics 2018, 84:11–16.

37. Gopalakrishnan K, Khaitan SK, Choudhary A, Agrawal A: Deep Convolutional Neural Networks with transfer learning for computer vision-based data-driven pavement distress detection. Construction and Building Materials 2017, 157:322–330.

38. Pan SJ, Yang Q: A survey on transfer learning. IEEE Transactions on knowledge and data engineering 2009, 22(10):1345–1359.

39. Wu EM, Wong LL, Hernandez BY, Ji JF, Jia W, Kwee SA, Kalathil S: Gender differences in hepatocellular cancer: disparities in nonalcoholic fatty liver disease/steatohepatitis and liver transplantation. Hepatoma research 2018, 4.

40. Li Y, Xu A, Jia S, Huang J: Recent advances in the molecular mechanism of sex disparity in hepatocellular carcinoma. Oncology letters 2019, 17(5):4222–4228.

41. Mauvais-Jarvis F, Bairey Merz N, Barnes PJ, Brinton RD, Carrero JJ, DeMeo DL, De Vries GJ, Epperson CN, Govindan R, Klein SL et al: Sex and gender: modifiers of health, disease, and medicine. Lancet 2020, 396(10250):565–582.

42. Costa AR, Lança de Oliveira M, Cruz I, Gonçalves I, Cascalheira JF, Santos CRA: The Sex Bias of Cancer. Trends in endocrinology and metabolism: TEM 2020, 31(10):785–799.

43. Pan JJ, Fallon MB: Gender and racial differences in nonalcoholic fatty liver disease. World journal of hepatology 2014, 6(5):274–283.

44. Bellamy RK, Dey K, Hind M, Hoffman SC, Houde S, Kannan K, Lohia P, Martino J, Mehta S, Mojsilovic A: AI Fairness 360: An extensible toolkit for detecting, understanding, and mitigating unwanted algorithmic bias. arXiv preprint arXiv:181001943 2018.

45. Schwab P, Mehrjou A, Parbhoo S, Celi LA, Hetzel J, Hofer M, Schölkopf B, Bauer S: Real-time prediction of COVID-19 related mortality using electronic health records. Nat Commun 2021, 12(1):1058.

