## Supplementary File for "Developing deep learning-based strategies to predict the risk of hepatocellular carcinoma among patients with nonalcoholic fatty liver disease from electronic health records"

### Supplementary Materials

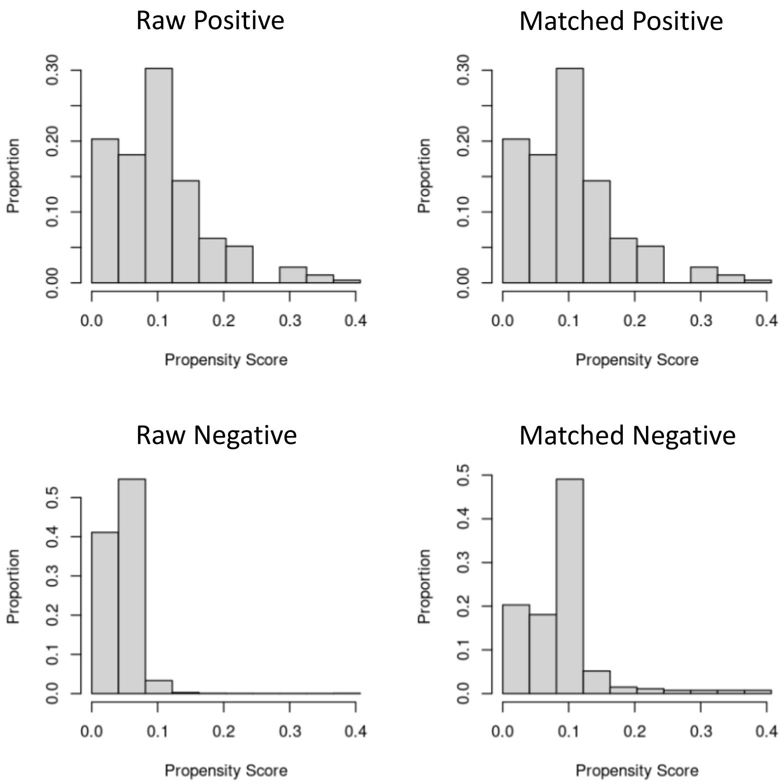


**Figure S1**. The distribution of propensity scores for positive and negative samples before and after propensity score matching. The distribution of the matched negative samples is closer to the positive samples.

### RETAIN model

### A

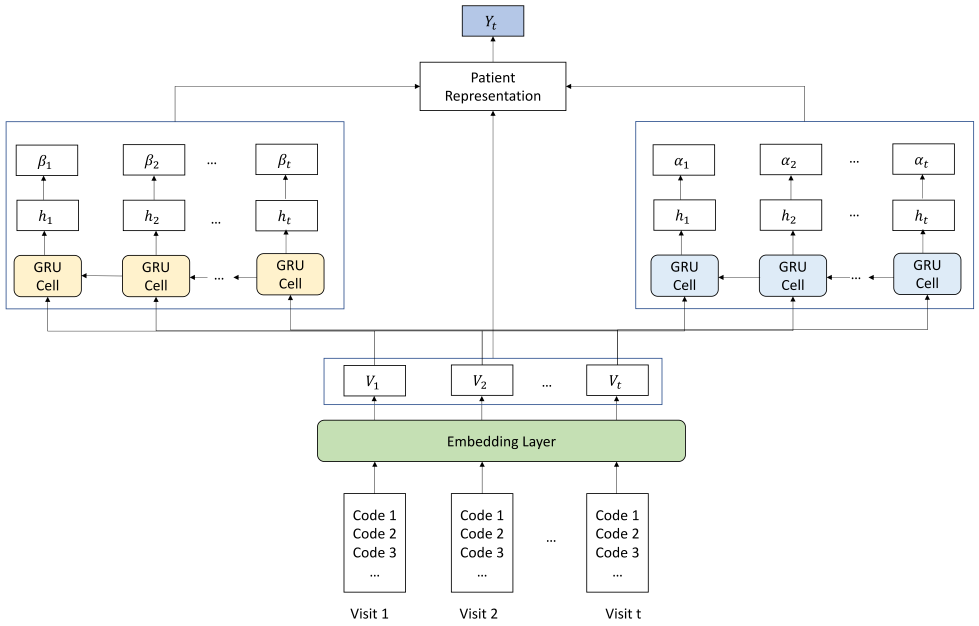


### B

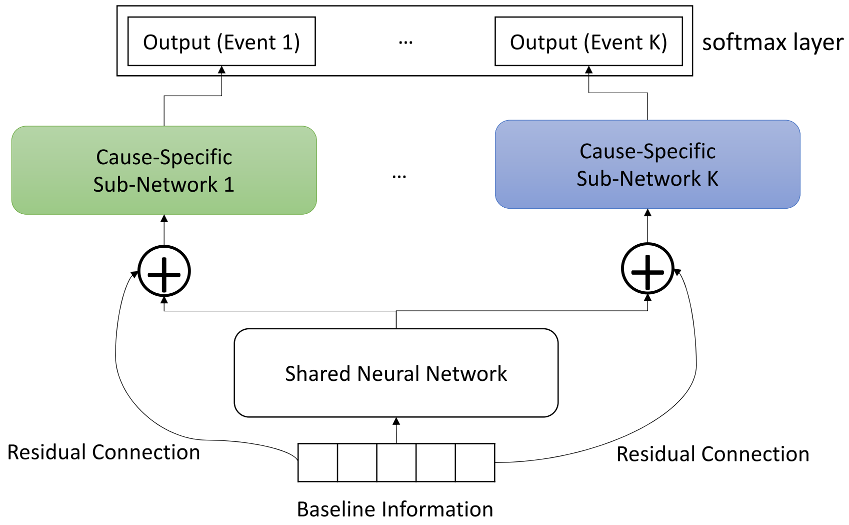


**Figure S2**. (A) The overview of RETAIN model. (B) The architecture of DeepHit model.

A common approach for analyzing time-to-event data with a classifier is to use the event status alone while ignoring the event times. Instead, we derive class labels by examining the event status at a chosen time point while removing patients who have insufficient follow-up (**Figure 2**). Specifically, the classification problem in this study is to predict NAFLD patients who will develop HCC within 10 years and those who will not, and the main objective is to identify risk factors and protective factors associated with the occurrence of HCC. We showed with Monte Carlo simulations that analyzing time-to-event data as a classification problem in this way allows us to identify risk factors reliably with strong control of type I error (Supplementary **Figure S1-2**). Further, under many conditions, the frequency of the positive class label is a well-calibrated estimator of the disease risk at the chosen time point (Supplementary **Figure S3**), although we do not need this property to identify risk factors. Accordingly, with this formulation, we can compare the performances of DeepHit and RETAIN in terms of predicting whether patients in a defined cohort develop a disease of interest within a specified time.

The formulation of disease prediction as a classification problem enabled us to take advantage of powerful deep learning models not developed for time-to-event data with censoring. We reframed the time-to-event problem as a classification problem by assigning class labels based on whether the event has occurred within a pre-specified time threshold, using the observed event times and statuses. We showed this formulation has desirable statistical properties in terms of the high accuracy of identifying risk factors and the calibration of the class probability as an estimator for the disease risk. This formulation thus allowed us to apply the RETAIN deep learning model in addition to the more statistically rigorous DeepHit model to identify risk factors for HCC progression among NAFLD patients, as well as exploring sex-specific patterns of HCC progression. It also makes it convenient to incorporate longitudinal information in the classification algorithm without specialized models for longitudinal data.

Our classification framework does not consider competing risks, so the identified risk factors must be interpreted carefully. For example, we showed that being a non-smoker appeared to be a risk factor for HCC in a NAFLD patient, which is inconsistent with a prior large retrospective study. A more reasonable interpretation would be that being a non-smoker lowers the competing risk of death and thus allows NAFLD patients more time to develop HCC. Moreover, to estimate disease risk accurately in this setting, we continue to advocate the use of competing risk models for now, especially those that can account for changes in covariate values over time. The situation could be handled by considering a new, multi-category classification algorithm in the future. Nonetheless, our formulation of disease prediction as a classification problem facilitates the application of powerful longitudinal deep learning models that do not model event censoring and thus provides timely clinical insights into disease progression over time. We expect that this algorithm works best in common situations where there are notable changes in a patient's health in the years close to the clinical event.

We showed that powerful predictive models such as deep learning can be sensitive to covariate imbalance, such as sex bias. The performance of disease prediction decreased by > 5% when RETAIN was trained data from on one sex and tested on data for the other sex. This performance reduction occurs because RETAIN can identify sex-specific features. For example, in female patients with NAFLD, analysis of the attention weights in the trained RETAIN models revealed that rheumatoid arthritis may be a risk factor and kidney stones may be a protective factor for HCC progression. Our results thus indicate that using a sex-biased dataset for training can reduce the predictive performance and generalizability of the trained deep learning model to other datasets. This finding can also be applied to other disease prediction tasks where sex and other patient characteristics such as race and ethnicity play important roles in disease progression.

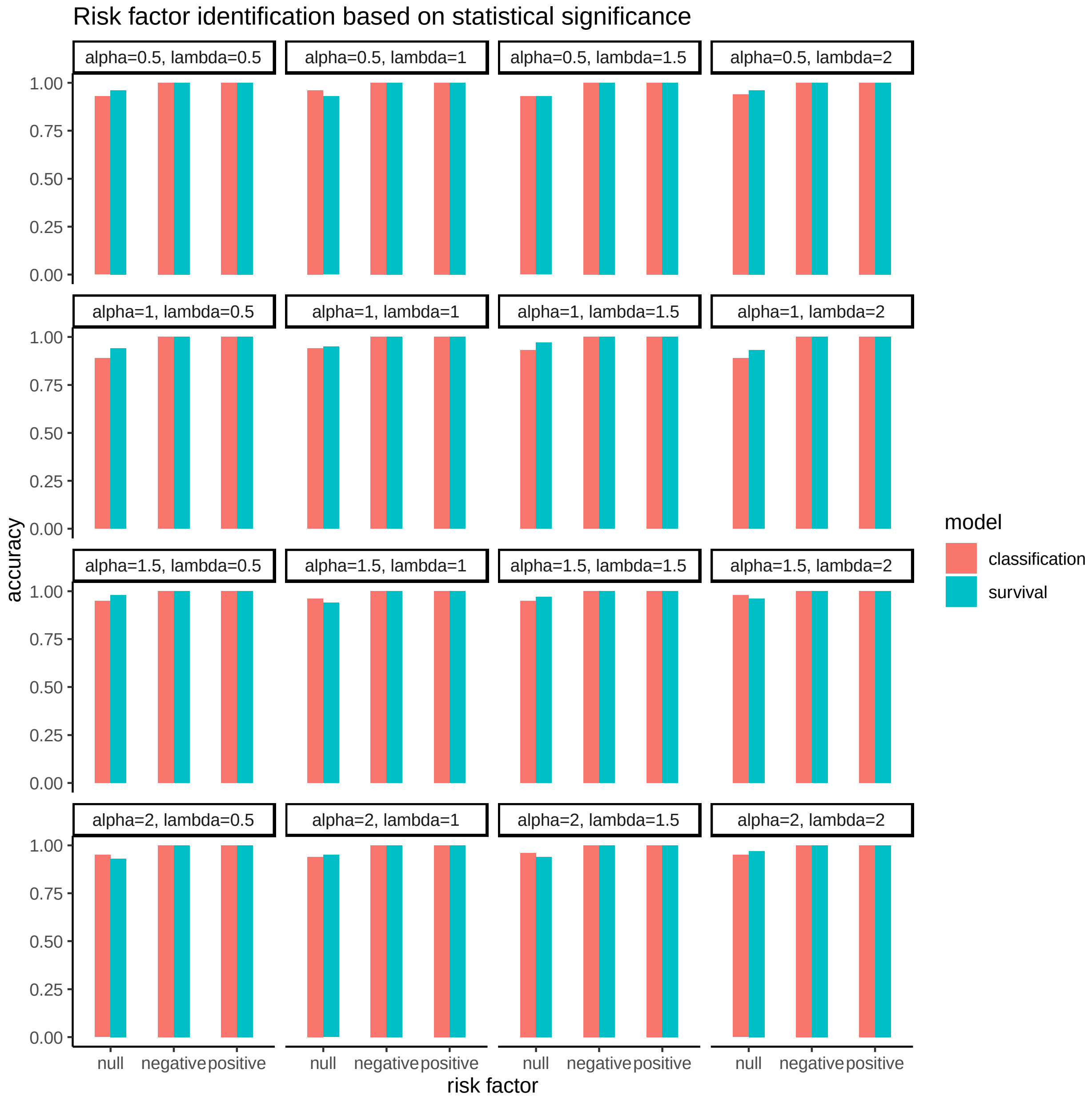


**Figure S3**. Comparison of the accuracy of risk factor identification under a survival model vs. a classification model. Each bar represents the accuracy of identifying a null, negative, or positive risk factor over 100 rounds of simulation based on whether the indicated model identified the risk factor as statistically significant. Time-to-event data (n = 1000) were generated with event times following the Weibull distribution with various shape parameters (alpha) and rate parameters (lambda), under a uniform censoring scheme. The hazard rates were modified multiplicatively by a null, negative, or positive risk factor. The time-to-event analysis used the Cox proportional-hazards regression to identify risk factors, while the classification analysis used the logistic regression for which class labels were defined as event occurrence within a time cutoff. Results are shown for the cutoff set at the 75% quantile of observed event times, and similar results were obtained for cutoff set at the 25% and 50% quantiles.

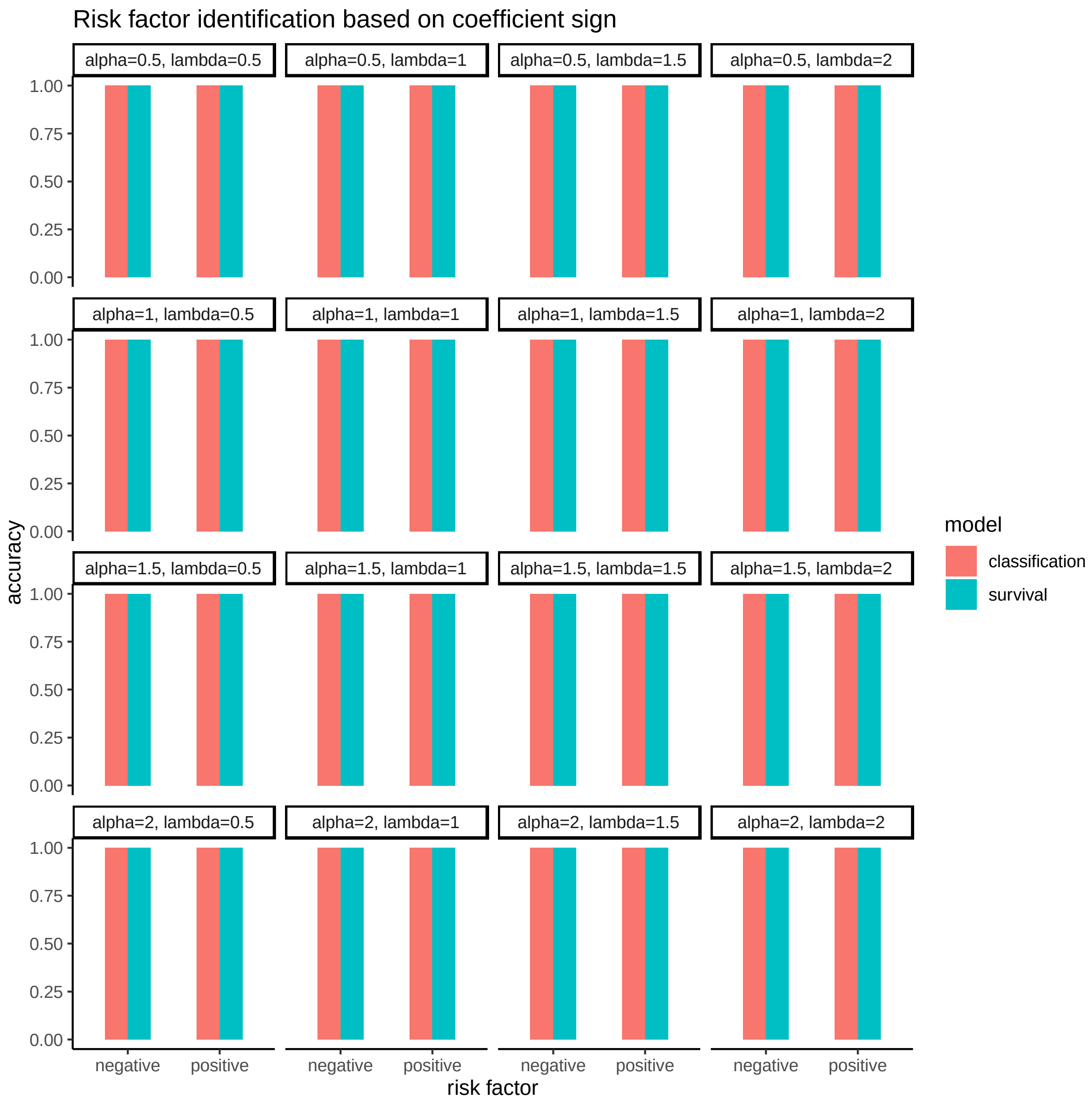


**Figure S4**. Comparison of the accuracy of risk factor identification under a survival model vs. a classification model, using the signs of the risk factor coefficient estimates. Each bar represents the accuracy of identifying a negative or positive risk factor over 100 rounds of simulation based on whether the indicated model estimated the coefficient for the risk factor with the correct sign (in the correct direction).

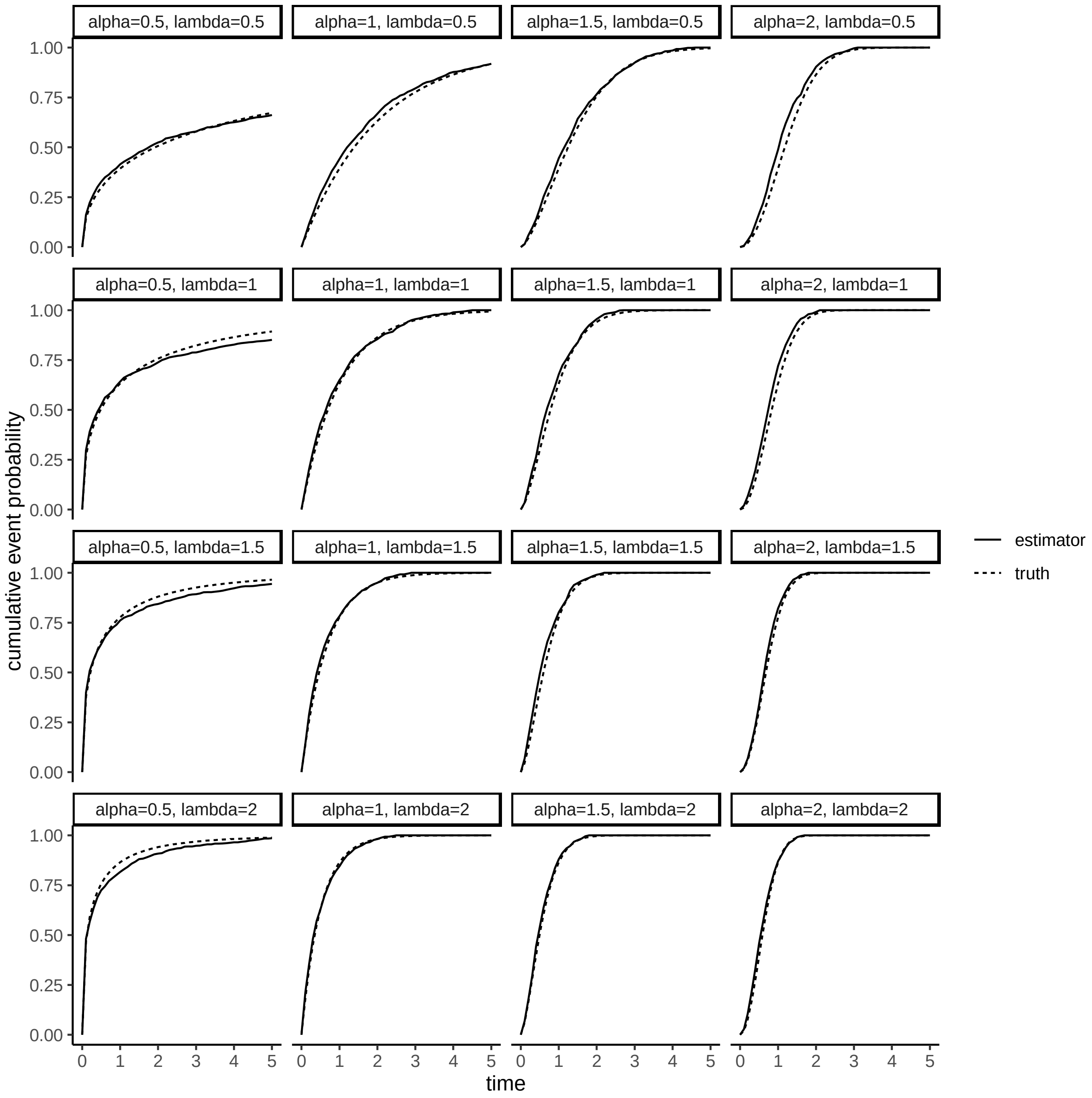


**Figure S5**. Class label probability is a well-calibrated estimator of the cumulative event probability under various conditions. Time-to-event data were generated using different Weibull distributions under a uniform censoring scheme. Class labels were defined based on whether the event was observed to occur before each time point.

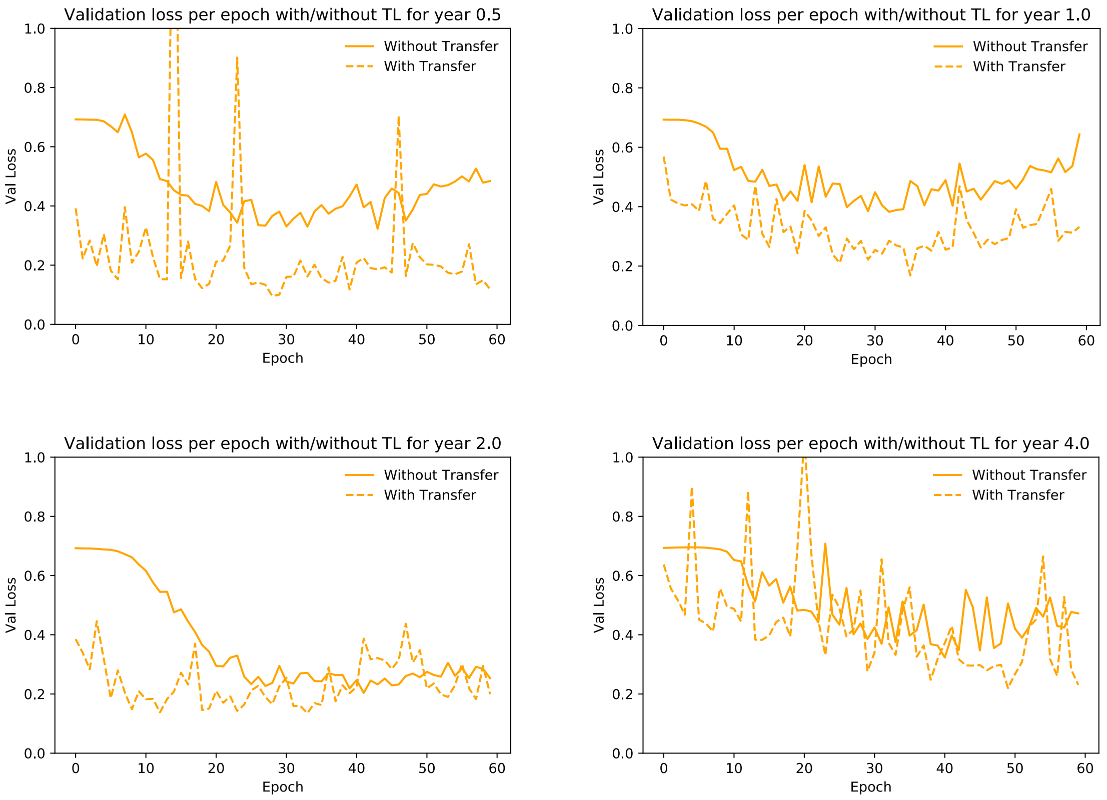


**Figure S6.** The validation loss with and without transfer learning under different masking length.

**Table S1**. Baseline variables for DeepHit.

| Name | Type |
|---|---|
| Marital status | multiple |
| Race | multiple |
| tobacco_use_index | binary |
| diabetes_index | binary |
| hypertension_index | binary |
| thrombocytopenia_index | binary |
| athero_index | binary |
| hypothyroidism_index | binary |
| CKD_index | binary |
| COP_index | binary |
| hyperlipidemia_index | binary |
| cirrhosis | binary |
| high_fib4 | binary |
| p_high_fib4 | binary |
| obesity | binary |

**Table S2**. High contribution medical codes with positive attention for male patients.

| rank | code | annotation |
|---|---|---|
| 0 | SMOKE_1 | Use tobacco |
| 1 | BMI_E | 35.0<bmi<40.0 |
| 2 | DIAG_Z51.89 | Encounter for other specified aftercare |
| 3 | MED_ONDANSETRON | |
| 4 | MED_PROPOFOL | |
| 5 | LAB_1920-8_High | Aspartate aminotransferase |
| 6 | MED_SODIUM CHLORIDE | |
| 7 | SMOKE_0 | No tobacco |
| 8 | MED_LVP SOLUTION | |
| 9 | DIAG_Z01.818 | Encounter for other preprocedural examination |
| 10 | MED_MORPHINE | |
| 11 | DIAG_I10 | Essential (primary) hypertension |
| 12 | DIAG_Z79.891 | Long term (current) use of opiate analgesic |
| 13 | DIAG_E11.9 | Type 2 diabetes mellitus without complications |
| 14 | MED_METOPROLOL | |
| 15 | BMI_ERROR | Null or negative |
| 16 | MED_ACETAMINOPHEN-OXYCODONE | |
| 17 | DIAG_F17.200 | Nicotine dependence unspecified uncomplicated |
| 18 | MED_DOCUSATE-SENNA | |
| 19 | DIAG_Z95.1 | Presence of aortocoronary bypass graft |

**Table S3**. High contribution medical codes with positive attention for female patients.

| rank | code | annotation |
|---|---|---|
| 0 | DIAG_M06.9 | Rheumatoid arthritis unspecified |
| 1 | DIAG_K76.89 | Other specified diseases of liver |
| 2 | DIAG_Z01.818 | Encounter for other preprocedural examination |
| 3 | BMI_E | 35.0<bmi<40.0 |
| 4 | SMOKE_0 | No tobacco |
| 5 | DIAG_E11.65 | Type 2 diabetes mellitus with hyperglycemia |
| 6 | LAB_1920-8_Low | Aspartate aminotransferase |
| 7 | SMOKE_1 | Use tobacco |
| 8 | DIAG_Z51.89 | Encounter for other specified aftercare |

| | | |
|---|---|---|
| 9 | DIAG_R19.7 | Diarrhea unspecified |
| 10 | GEND_female | |
| 11 | MED_ONDANSETRON | |
| 12 | MED_SODIUM CHLORIDE | |
| 13 | DIAG_I10 | Essential (primary) hypertension |
| 14 | LAB_1920-8_High | Aspartate aminotransferase |
| 15 | DIAG_R42 | Dizziness and giddiness |
| 16 | DIAG_R05 | Cough |
| 17 | DIAG_C78.7 | Secondary malignant neoplasm of liver and intr... |
| 18 | BMI_C | 25.0<bmi<30.0 |
| 19 | MED_INSULIN GLARGINE | |

**Table S4**. High contribution medical codes with negative attention for male patients.

| **rank** | **code** | **annotation** |
|---|---|---|
| 0 | LAB_1920-8_In Control | Aspartate aminotransferase |
| 1 | MED_ZOLPIDEM | |
| 2 | MED_KETOROLAC | |
| 3 | A_J | Age_50 |
| 4 | MED_DIPHENHYDRAMINE | |
| 5 | MED_ATORVASTATIN | |
| 6 | A_H | Age_40 |
| 7 | MED_EPTIFIBATIDE | |
| 8 | A_I | Age_45 |
| 9 | MED_MAGNESIUM HYDROXIDE | |
| 10 | MED_HYDROMORPHONE | |
| 11 | LAB_777-3_Within Range | Platelets |
| 12 | MED_ACETAMINOPHEN | |
| 13 | MARI_null | |
| 14 | MED_PANTOPRAZOLE | |
| 15 | MED_DOCUSATE | |
| 16 | DIAG_I20.0 | Unstable angina |
| 17 | MED_STERILE WATER | |
| 18 | MED_AMLODIPINE | |
| 19 | MED_METOCLOPRAMIDE | |

**Table S5**. High contribution medical codes with negative attention for female patients.

| rank | code | annotation |
|---|---|---|
| 0 | DIAG_N20.0 | Calculus of kidney |
| 1 | A_I | Age_45 |
| 2 | MED_CIPROFLOXACIN | |
| 3 | A_J | Age_50 |
| 4 | MED_PANTOPRAZOLE | |
| 5 | MED_KETOROLAC | |
| 6 | MED_AMLODIPINE | |
| 7 | MED_HYDROMORPHONE | |
| 8 | MED_STERILE WATER | |
| 9 | MED_LEVOFLOXACIN | |
| 10 | MED_DIPHENHYDRAMINE | |
| 11 | MED_CLINDAMYCIN | |
| 12 | A_K | Age_55 |
| 13 | LAB_777-3_Within Range | Platelets |
| 14 | MED_DOCUSATE | |
| 15 | MED_ATORVASTATIN | |
| 16 | MED_METRONIDAZOLE | |
| 17 | MARI_null | |
| 18 | MED_ACETAMINOPHEN | |
| 19 | LAB_777-3_Low | Platelets |

**Inclusion and Exclusion Criteria for Non-alcoholic fatty liver disease (NAFLD) cohort from EHR**

*Inclusion:*

1) patients were classified as having NAFLD if they had 2 or more elevated alanine aminotransferase (ALT) values (≥40 IU/mL for men and ≥31 IU/mL for women) in the ambulatory settings and more than 6 months apart, with no positive serologic testing for HBV (ie, HBV surface antigen) or HCV (ie, HCV RNA);

2) ≥18 years old at index date of follow-up (the date of first elevated ALT as the index date of follow-up for NAFLD cases).

*Exclusion:*

1) alcohol-related ICD-9 codes any time before or during study follow-up, ICD9: 571.0 Alcoholic fatty liver; 571.1 Acute alcoholic hepatitis; 571.2 Alcoholic cirrhosis of liver;571.3 Alcoholic liver damage, unspecified. ICD10: K70.0 Alcoholic fatty liver; K70.10 Alcoholic hepatitis without ascites; K70.30 Alcoholic cirrhosis of liver without ascites; K70.9 Alcoholic liver disease, unspecified;

2) positive AUDIT-C scores (≥4 in men and ≥3 in women) any time before or during study follow-up;

3) evidence of rare chronic hepatitides (eg, hereditary hemochromatosis, primary biliary cirrhosis, primary sclersoing cholangitis, a-1 antitrypsin disease, or autoimmune hepatitis) based on ICD-9 codes, ICD9: 713.0 Arthropathy associated with other endocrine and metabolic disorders (Code first underlying disease, as hemochromatosis [275.0]); 571.6 Biliary cirrhosis; 576.1 Cholangitis; 273.4 Alpha-1-antitrypsin deficiency; 571.42 Autoimmune hepatitis. ICD10: M14.80 Arthropathies in other specified diseases classified elsewhere, unspecified site; K74.3 Primary biliary cirrhosis; K83.0 Cholangitis; E88.01 Alpha-1-antitrypsin deficiency; K75.4 Autoimmune hepatitis.

**Control cohort**

*Inclusion:*

1) an ALT test performed;

2) no any documented liver-related risk factor: no NAFLD (persistently normal ALT); absence of positive tests for HBV and HCV; absence of alcohol ICD codes, ICD9: 571.0 Alcoholic fatty liver; 571.1 Acute alcoholic hepatitis; 571.2 Alcoholic cirrhosis of liver;571.3 Alcoholic liver

damage, unspecified, ICD10: K70.0 Alcoholic fatty liver; K70.10 Alcoholic hepatitis without ascites; K70.30 Alcoholic cirrhosis of liver without ascites; K70.9 Alcoholic liver disease, unspecified;

3) all AUDIT-C scores (<4 in men and <3 in women);

4) the date of first ALT test in the study timeframe as the index date of follow-up for controls;

5) random sampling without replacement (case: control= 1:1): sex; age at first ALT (index date); duration from their first visit to the first ALT test date (These three conditions of case are best the same as the control. If it is not the same, we can take the nearest one.).

**Inclusion and Exclusion Criteria for Hepatocellular cancer (HCC) cohort from EHR**

*Inclusion:*

1) patients diagnosed with HCC (4 ICD codes for HCC: 155.0, C22.0, C22.8 and C22.9)

2) ≥18 years old at first visit date

*Exclusion:*

1) patients were classified as having non-alcoholic fatty liver disease (NAFLD) if they had 2 or more elevated alanine aminotransferase (ALT) values (≥40 IU/mL for men and ≥31 IU/mL for women) in the ambulatory settings and more than 6 months apart (loinc code for ALT test: 1742-6)

**Control cohort**

*Inclusion:*

1) patients diagnosed without liver cancer (14 ICD codes for liver cancer: 155, 155.0, 155.1, 155.2, 197.7, 209.72, C22.0, C22.1, C22.2, C22.7, C22.8, C22.9, C78.7 and C7B.02)

2) ≥18 years old at first visit date

3) random sampling without replacement (case: control= 1:10): same gender; same age at first visit date; same duration from the first visit to the last visit date (If the sample size of the exact matched controls is not enough, please use the nearest to match).

*Exclusion:*

1) patients were classified as having NAFLD if they had 2 or more elevated alanine aminotransferase (ALT) values (≥40 IU/mL for men and ≥31 IU/mL for women) in the ambulatory settings and more than 6 months apart (loinc code for ALT test: 1742-6)
